# Understanding eye care needs beyond vision loss across six countries: addressing distance and near visual impairment and other eye conditions

**DOI:** 10.64898/2026.09.01.26360563

**Authors:** Andrew Bastawrous, Marzieh Katibeh, Sergio Latorre-Arteaga, Michael Gichangi, Asad Khan, Shalinder Sabherwal, Moses Kasadhakawo, Sailesh Mishra, Rebecca Oenga, Furahini Godfrey Mndeme, Allen Foster

## Abstract

**Objective:** Population growth and ageing are placing increasing pressure on eye care services. We present a model estimating comprehensive eye care needs using data across six countries.

**Methods and analysis:** We used cross-sectional data from community screening programmes (n = 2,338,193; all ages) conducted in 2022–2023 in India, Pakistan and Nepal (South Asia) and Kenya, Tanzania and Uganda (sub-Saharan Africa). Total eye care need was defined as the presence of distance visual impairment (VI), near VI, or any other eye condition requiring care in at least one eye, reported with 95% confidence intervals (CI). Distance and near VI were assessed on presenting visual acuity, measured with existing correction where available; estimates therefore reflect unmet need rather than total impairment. Logistic regression identified factors associated with increased need, reported as adjusted odds ratios (AORs).

**Results:** Among those aged ≥50 years, the prevalence of distance VI was 36.3% (95% CI: 36.1– 36.5), and 74.8% (95% CI: 74.6–74.9) had at least one eye care need. In those aged <50 years, the prevalence of distance VI was 6.3% (95% CI: 6.2–6.3), and 19.9% (95% CI: 19.8–19.9) had at least one eye care need. Higher need was observed among older individuals (AOR: 1.06 per year), females (AOR: 1.13), and in programmes conducted in sub-Saharan Africa compared to South Asia (AOR: 2.75); all p-values < 0.001.

**Conclusion:** This study expands knowledge by including all age groups, all types of eye care needs, and each affected eye, not only bilateral problems, offering a more complete picture of eye care needs in mass screening programmes.

**What is already known on this topic:** Most eye care survey data before 2024 focus on distance visual impairment and its causes in people aged 50+. Near vision impairment and other eye conditions are often excluded from population surveys due to feasibility constraints.

**What this study adds:** As a result, current estimates likely underestimate the true burden of eye care needs. Analysis of >2.3 million people screened in six countries provides a more comprehensive estimate of population eye care needs.

**How this study might affect research, practice or policy:** These results highlight the importance of planning services for a wider range of eye care needs, including near vision correction and other conditions. In addition, this study demonstrates the potential of large-scale digital programme data to inform population eye health monitoring and service planning.

## Introduction

Improving eye health is a practical and cost-effective way of unlocking human potential. [1] Addressing vision impairment is therefore central to achieving broader health and development goals, including universal health coverage and the Sustainable Development Goals. [2] Bibliometric analyses of global eye health research have highlighted substantial inequities in data generation, with a relative scarcity of evidence from low- and middle-income countries, particularly in sub-Saharan Africa and South Asia where the burden is greatest. [3]

Population-based surveys, such as the Rapid Assessment of Avoidable Blindness (RAAB), have been instrumental in providing representative estimates of vision impairment and informing national planning. [4,5] These surveys are designed to be practical and cost-effective, often focusing on populations aged 50 years and above, where the burden of blindness is highest. In addition, by design, most eye care survey data before 2024 do not include near vision loss or non-vision-impairing eye conditions. [5]

In parallel, early identification of eye care needs through community and primary care screening enables timely treatment and management, particularly in settings with limited access to facility-based services. [6–8] While such programme data are not inherently population-representative, when collected systematically and at scale they offer valuable, real-world insights into eye care needs across all age groups, including near vision impairment and a broader range of ocular conditions. [9]

In this analysis, we used data generated from eye health programmes in regions in sub-Saharan Africa and South Asia using Peek, an end-to-end software and data insights platform designed for eye health. Datasets were selected where concurrent or closely timed surveys and community-based (door-to-door) screening data were available. [10]

The objective of this study was to quantify the overall burden of eye care needs using systematically collected community screening data, encompassing all ages and including distance and near vision impairment, as well as an estimate of other eye conditions requiring care.

## Methods

### Study design and sources of data

This study presents a cross-sectional analysis of eye health screening programmes conducted across six countries between January 2022 and September 2023 using the Peek platform. Approximately 80% of participants were enrolled during door-to-door visits by community health workers, with the remainder 20% enrolled at primary health facilities in the community eye care programmes.

Sample size included 2,338,193 individuals. Among them, 109,307 (4.7%) were not tested for distance vision, primarily children under five or adults with cognitive difficulties. However, all children were screened for other eye conditions, and those with suspected visual problems were referred for further assessment.

### Inclusion and exclusion criteria

All individuals who presented at the household and were willing to participate in the screening programmes were included in the analysis. There were no predefined exclusion criteria, no age restrictions were applied.

Among the participants, 58.6% were female, 41.3% were male, and 0.03% (n = 726) identified as transgender. A total of 83.8% were younger than 50 years, and 16.2% were 50 years or older.

### Outcome definitions

Eye care needs across all ages were categorised into three main groups: distance visual impairment (VI), near VI, and other eye conditions. The definition of distance VI was identified by presenting visual acuity (PVA) measured with existing correction if available. VI was defined according to the World Health Organization / International Classification of Disease-11 (ICD-11) definitions as PVA from less than 6/12 in adults and less than 6/9 in children. [8]

Near VI was defined as binocular presenting near VA less than N8 with existing correction if available. In India, near VI was subjectively assessed based on patients’ perceptions of difficulties with near vision. Following distance and near vision tests, external eye examinations were conducted by trained screening cadre to identify patients with other eye conditions, such as red eyes, pain, external lesions, and to gather information on conditions such as diabetes and glaucoma.

### Patient and public involvement

Local health authorities and primary healthcare representatives were involved in the design and implementation of the programmes and referral pathways. Patients were not involved in the design, conduct, reporting, or dissemination plans of this research.

### Ethical considerations

The screening programmes were delivered as part of routine eye care service delivery, with the involvement of local health authorities, and participation was voluntary. Review by a research ethics committee was not required, as the data used in this study were routine programme data collected as part of service delivery, and the analysis was conducted on anonymised data. Data storage, transmission and retrieval were governed by Data Protection Agreements with local stakeholders in each country and complied with the European Union General Data Protection Regulation (GDPR).

### Statistical analysis

Data were analysed using Stata version 14.0 (StataCorp, College Station, TX, USA). Descriptive analyses were conducted to summarise eye care needs by age, sex, and programme location. Programme locations were grouped into two broad regions: South Asia (India, Pakistan, and Nepal) and sub-Saharan Africa (Kenya, Tanzania, and Uganda). Logistic regression was used to identify factors associated with increased eye care need, and results were reported as adjusted odds ratios (AORs) with corresponding confidence intervals.

## Results

In the total screened population in both regions among individuals aged over 50 years, the prevalence of distance VI in at least one eye was 36.3%. When including unmet needs related to near VI and other eye conditions identified through the screening programmes, an estimated 74.8% of individuals aged over 50 years had at least one eye care need. Among those under 50 years, the prevalence of distance VI and any eye care needs in at least one eye was 6.3% and 19.9%, respectively.

Table 1 presents the prevalence of distance VI in at least one eye and the overall eye care needs in two broad age groups in the screening programmes across six countries.

**Table 1.** Estimation of distance visual impairment (VI) and the overall eye care needs in the programmes conducted in the two global regions.

| Country / region | Age 0–49 years |  | Age 50+ years |  |
| --- | --- | --- | --- | --- |
|  | Distance VI | Any need | Distance VI | Any need |
|  | % (95% CI) | % (95% CI) | % (95% CI) | % (95% CI) |
| India | 2.5 (2.4–2.6) | 10.7 (10.6–10.9) | 42.8 (42.2–43.3) | 61.1 (60.6–61.6) |
| Nepal | 4.2 (3.9–4.4) | 14.1 (13.7–14.5) | 34.3 (33.3–35.4) | 56.7 (55.6–57.7) |
| Pakistan | 6.2 (6.2–6.3) | 11.5 (11.4–11.6) | 36.1 (35.6–36.5) | 52.6 (52.1–53.0) |
| **Average SA** | **5.5 (5.4–5.6)** | **11.4 (11.4–11.5)** | **38.3 (38.0–38.6)** | **56.0 (55.6–56.3)** |
| Kenya | 6.9 (6.8–7.1) | 27.4 (27.3–27.5) | 35.9 (35.6–36.0) | 81.5 (81.3–81.6) |
| Tanzania | 8.1 (7.8–8.2) | 31.5 (31.2–31.8) | 28.4 (27.5–29.3) | 57.1 (56.2–58.1) |
| Uganda | 10.4 (9.9–10.9) | 52.9 (52.1–53.6) | 42.7 (41.2–44.1) | 89.5 (88.6–90.4) |
| **Average SSA** | **7.1 (7.0–7.1)** | **28.1 (28.0–28.2)** | **35.7 (35.5–35.9)** | **80.7 (80.6–80.9)** |

Data presented in the table are weighted averages. Distance VI: presenting visual acuity in at least one eye $< 6/12$ in adults and $< 6/9$ in children. Any need: distance VI, near VI, and/or other eye conditions in at least one eye. SA: South Asia; SSA: sub-Saharan Africa.

Figure 1 presents the prevalence of unmet eye care needs by demographic characteristics, including age, sex, and screening location.

**Figure 1.**
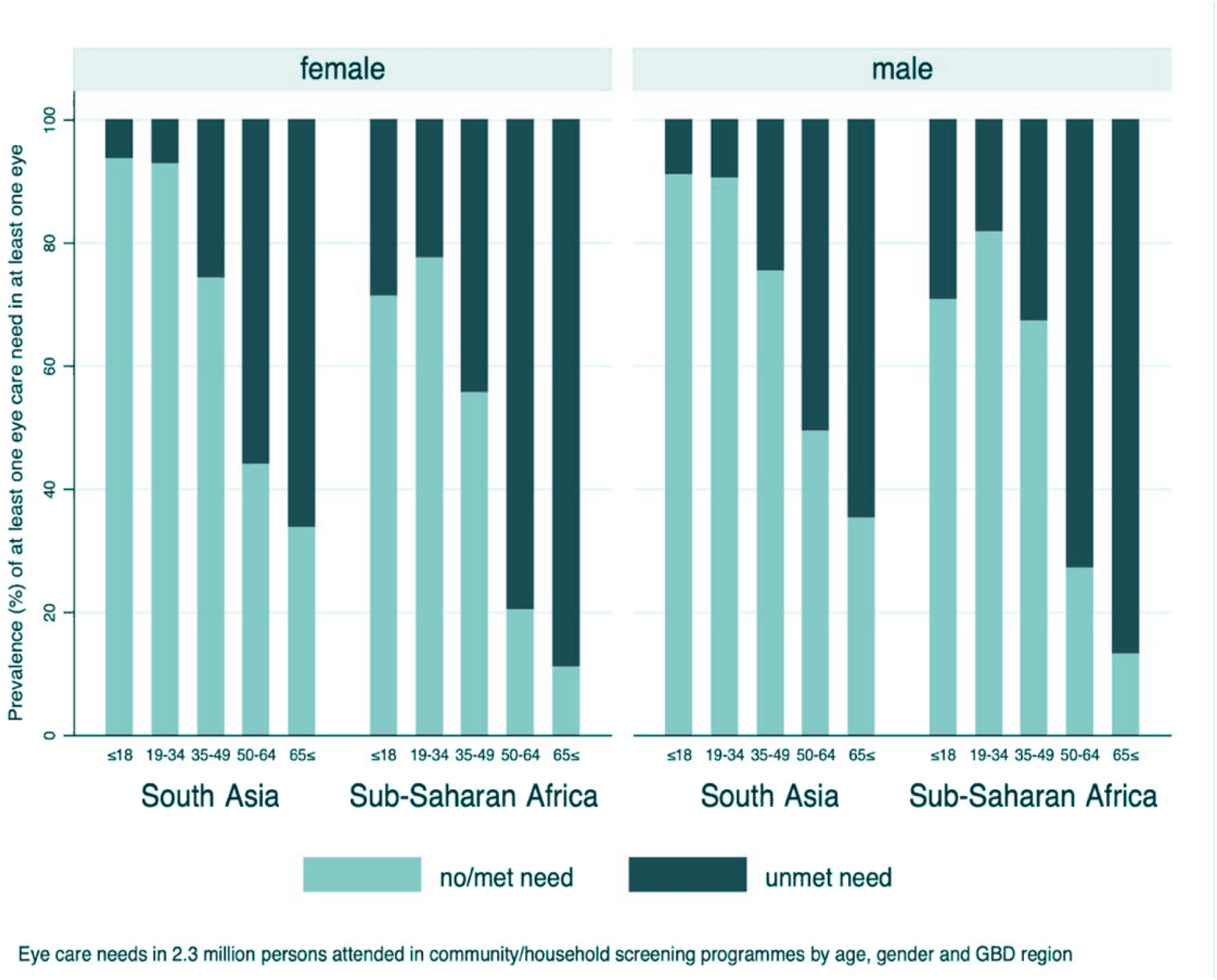
Distribution of eye care needs based on community-based screening programmes across six countries, categorised by age, sex, and region

Figure 2 illustrates the overlap of different eye care needs among the total screened population. Notably, this figure highlights a subgroup with isolated near vision impairment who may benefit from ready-made reading spectacles, which could be provided at lower levels of the eye care referral system.

**Figure 2.**
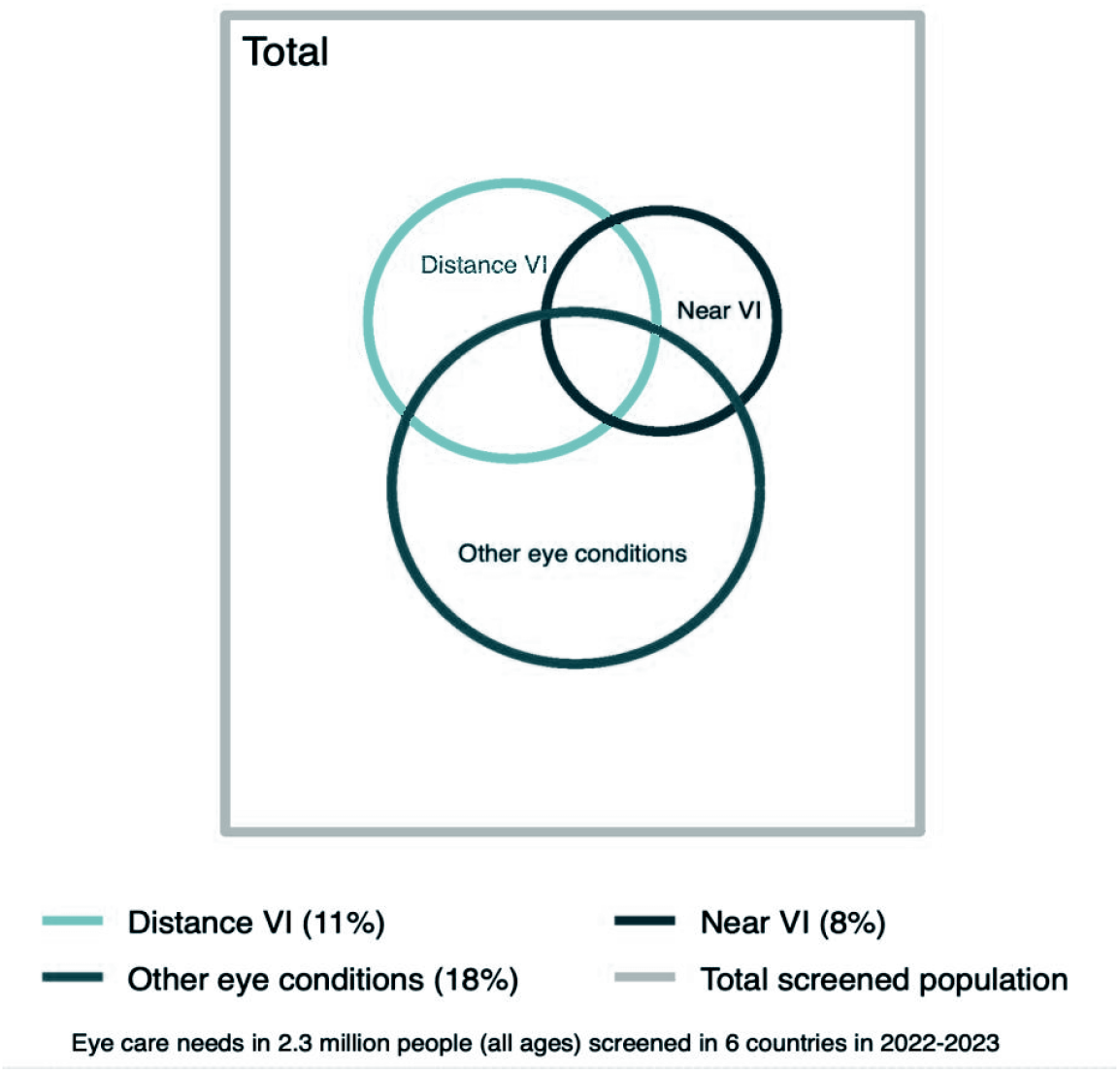
Illustration of eye care needs from 2.3 million participants of all age groups screened through door-to-door and community-based programmes across six countries

Analysis of adjusted odds ratios (AOR) showed that the need for eye care increased with age (AOR for each year of ageing: 1.06, 95% CI: 1.061–1.062), female sex (AOR: 1.13, 95% CI: 1.12– 1.14), and programmes in sub-Saharan Africa (AOR: 2.75, 95% CI: 2.73–2.77).

## Discussion

Hundreds of millions of people are affected by preventable or treatable poor vision, which affects their education, employment, productivity, wellbeing, and safety. [11] The data in this manuscript highlight a substantial demand for eye care services across all age groups.

The leading causes of vision (distance and/or near) impairment (VI), namely cataracts, myopia, and presbyopia, account for close to 1 billion cases of blindness or VI worldwide. [12] All of these conditions have proven, cost-effective treatments, although many people do not access services due to limited resources, insufficient eye care planning, and a lack of awareness of patients and health care providers. [3]

Community-based screening programmes are increasingly implemented to identify various eye care needs and link individuals to services across all age groups. [13,14] Drawing from data obtained from 2.3 million people screened through door-to-door or community health units, our data show approximately 3 in 4 people over the age of 50 and 1 in 5 people under the age of 50 have at least one eye care need, that requires referral or management at the screening site, for instance near vision impairment due to presbyopia.

Based on data from the UN’s World Population Prospects, [15] 5.1% of the African population and 11.9% of the population in South Asia are aged 50 years or above. By applying these proportions to the eye care needs in the programme data (Table 1), the estimates of eye care needs are presented in Box 1. These estimates should not be interpreted as precise prevalence measures, but rather as illustrative projections to support service planning.

#### Box 1. Estimation of eye care needs for screening programmes of all ages

**For a district in sub-Saharan Africa**

(a) 86 people/1000 total population have a distance VI (in either eye)
(b) 78 people/1000 total population have near VI
(c) 232 people/1000 total population have all other eye conditions **308 people/1000 total population have any eye care needs (a, b or c)**

**For a district in South Asia**

(a) 94 people/1000 total population have distance VI (in either eye)
(b) 59 people/1000 total population have near VI
(c) 102 people/1000 total population have all other eye conditions **167 people/1000 total population have any eye care needs (a, b or c)**

The resulting estimates suggest that a substantial proportion of the total population, not only older adults, may require some form of eye care. In both regions, the burden extends beyond distance vision impairment to include near vision impairment and a large proportion of other eye conditions that may not be captured in traditional survey approaches. This aligns with previous evidence that a significant share of unmet eye care need relates to refractive error and conditions that affect functional vision but may not meet conventional thresholds for visual impairment. [2,12]

The higher estimated burden of eye care needs observed in sub-Saharan Africa compared with South Asia may reflect a combination of health system, epidemiological, and access-related factors. Sub-Saharan Africa continues to have some of the lowest levels of eye care workforce and infrastructure globally, with limited availability of services outside urban centres and very low specialist-to-population ratios. [16,17] As a result, access to effective interventions such as cataract surgery remains constrained, despite cataract being the leading cause of blindness. [1,12]

More broadly, vision impairment is strongly associated with socioeconomic disadvantage, rural residence, and barriers to accessing care, all of which are more pronounced in many sub-Saharan African settings. [2]

### Holistic approach

Historically, “avoidable blindness” served as a powerful focus and way of prioritising resources. The global agenda then expanded to include varying levels of distance vision impairment beyond blindness, and only very recently have global estimates included near vision loss. [16]

Community screening programme data have demonstrated that patients access eye care services for a combination of reasons, broadly: (a) distance VI, e.g. cataracts or myopia; (b) near VI, e.g. presbyopia; (c) all other eye conditions, e.g. conjunctivitis, or conditions that lead to risk of future vision loss, e.g. glaucoma. These three broad categories are not mutually exclusive, so patients can present with any individual category or a combination. Our findings show that drawing data from multiple sources—including across age groups and from door-to-door / community screening programmes—provides a holistic picture of the unmet eye health needs of a region. Combining all of these data helps plan resources and reduce inequity by providing programme planners with a better understanding of the specific needs in their region.

### Strengths and limitations

The estimates are based on programme data combined from several countries and reflect mass screening programmes rather than nationally representative prevalence data. Therefore, differences in sampling approaches and programme coverage across countries should be considered when interpreting these findings.

Despite this limitation, the use of large-scale, real-world programme data provides valuable insights into the broader spectrum of “all eye care needs” in diverse settings. These findings are relevant for informing the design and planning of screening programmes in similar contexts. The data also support the need to strengthen primary eye care and improve integration across outreach services, primary care, optometry, and ophthalmic services, including the development of effective referral pathways to ensure access to eye care for all.

## Conclusion

The projected estimates suggest that a considerable proportion of the population—far beyond those with distance vision impairment alone—may require some form of eye care, with needs spanning refractive services, management of non-blinding conditions, and referral for more complex care. These findings highlight the importance of designing eye care surveys and services that reflect the full spectrum of population needs across all age groups. Incorporating systematically collected programme data into planning processes can support more accurate estimation of service demand, enabling programme managers to better plan workforce requirements, allocate supplies, and strengthen referral pathways. Such approaches may contribute to more efficient, equitable, and people-centred eye care delivery in low-resource settings.

## Data Availability

All data produced in the present study are available upon reasonable request to the authors

## Declarations

### Ethics approval

The screening programmes were delivered as part of routine eye care service delivery, with the involvement of local health authorities, and participation was voluntary. Review by a research ethics committee was not required, as the data used in this study were routine programme data collected as part of service delivery, and the analysis was conducted on anonymised data.

### Data availability

The data underlying this study are routine programme data collected through eye health programmes using the Peek platform. Data storage, transmission and retrieval were governed by Data Protection Agreements with local stakeholders in each country and complied with the European Union General Data Protection Regulation. De-identified aggregate data may be available from the corresponding author on reasonable request, subject to approval from the relevant national programme partners.

